# THE INFLAMMATORY MICROENVIRONMENT IN SCREEN-DETECTED PREMALIGANT ADENOMATOUS POLYPS: EARLY RESULTS FROM THE INTEGRATED TECHNOLOGIES FOR IMPROVED POLYP SURVEILLANCE (INCISE) PROJECT

**DOI:** 10.1101/2020.08.16.20175935

**Authors:** David Mansouri, Stephen T McSorley, James H Park, Clare Orange, Paul G Horgan, Donald C McMillan, Joanne Edwards on behalf of the Integrated Technologies for Improved Polyp Surveillance (INCISE) collaborators

**Author notes:** Corresponding Author Mr David Mansouri, Consultant Colorectal Surgeon Glasgow Royal Infirmary, Glasgow, UK, G31 2ER. Disclosures: none. Funding: The Cunningham Trust, SCO13499, Ref ACC/KWF/CT12/21.

## Abstract

**Introduction:** Around 40% of patients who attend for colonoscopy following a positive stool screening test have adenomatous polyps. Identifying which patients have a higher propensity for malignant transformation is currently poorly understood. The aim of the present study was to assess whether the type and intensity of inflammatory infiltrate differs between high-grade (HGD) and low-grade dysplastic (LGD) screen detected adenomas.

**Methods:** A representative sample of 207 polyps from 134 individuals were included from a database of all patients with adenomas detected through the first round of the Scottish Bowel Screening Programme (SBoSP) in NHS GG&C (April 2009 to April 2011).

Inflammatory cell phenotype infiltrate was assessed by immunohistochemistry for CD3+, CD8+, CD45+ and CD68+ in a semi-quantitative manner at 20x resolution. Immune-cell infiltrate was graded as absent, weak, moderate or strong.

Patient and polyp characteristics and inflammatory infiltrate were then compared between HGD and LGD polyps.

**Results:** CD3+ infiltrate was significantly higher in HGD polyps compared to LGD polyps (74% vs 69%, p<0.05). CD8+ infiltrate was significantly higher in HGD polyps compared to LGD polyps (36% vs 13%, p<0.001) where as CD45+ infiltrate was not significantly different(69% vs 64%, p=0.401). There was no significant difference in CD68+ infiltrate (p=0.540) or total inflammatory cell infiltrate (calculated from CD3+ and CD68+) (p=0.226).

**Conclusions:** This study reports an increase in CD3+ and CD8+ infiltrate with progression from LGD to HGD in colonic adenomas. It may therefore have a use in the prognostic stratification and treatment of dysplastic polyps.

## Introduction

Screening for colorectal cancer utilising either the guaiac-based faecal occult blood test (gFOBt) or the faecal immunochemical test (FIT) has been shown to reduce cancer specific mortality through the detection of early stage disease (Mandel, Bond et al. 1993; Hardcastle, Chamberlain et al. 1996; Kronborg, Fenger et al. 1996). However, the majority of individuals who attend for colonoscopy following a positive screening test do not have cancer detected, but a large proportion do have adenomatous polyps. There is good evidence that colorectal cancer develops through the adenoma-carcinoma sequence and it has been estimated that approximately 25% of polyps greater than 1cm will develop into cancer over 20 years (Stryker, Wolff et al. 1987). There is some evidence from gFOBt screening that in the context of a high positivity rate the incidence of cancer in a given population can be reduced by removal of these polyps (Mandel, Church et al. 2000).

There is now a wealth of evidence that progression and outcome of colorectal cancer is related to a complex interaction between tumour and host (Hanahan and Weinberg 2011). In particular, those with a more pronounced peri-tumoural inflammatory reaction have better cancer specific outcomes (Roxburgh and McMillan 2012). However, it is not clear whether such a relationship is relevant to malignant adenomatous polyps. Previous studies examining this phenomenon have been limited by both numbers and a focus on early invasive cancer and have failed to exam the host inflammatory response across the spectrum of dysplasia. Therefore, our understanding of the natural history of such polyps and in particularly the role of the inflammatory infiltrate is limited (Cui, Yuan et al. 2009; McLean, Murray et al. 2011; Cui, Shi et al. 2012).

Despite such insights, determining which patients with polyps have a higher propensity for malignant transformation remains a challenge. Current UK guidelines advise repeat or surveillance colonoscopies depending on the number, size and grade of polyp detected at index procedure (Rutter et al. 2020). Nevertheless, there is limited evidence that this impacts on patient outcome, especially in those of low to intermediate risk (Cross et al. 2020). In addition, a recent population study has suggested that colorectal cancer mortality may actually be higher in those patients who have a high risk polyp removed compared with the general population (Loberg, Kalager et al. 2014).

Therefore, the goal of the Integrated Technologies for Improved Polyp Surveillance (INCISE) project is to combine polyp and tissue characteristics, in particular genomic, transcriptomic, and local inflammatory features which are associated with a higher likelihood of future polyps. This will allow for post-polypectomy surveillance with greater precision, allowing fewer unnecessary procedures for those at low risk, and concentrating valuable resource on those at high risk.

As part of this overall project, the aim of the present study was to assess the role of the local inflammatory response in screen-detected dysplastic adenomas and to assess whether the type and intensity of inflammatory infiltrate differs between high-grade and low-grade dysplasia.

## Materials and Methods

A database of all patients with adenomas detected through the first round of the Scottish Bowel Screening Programme (SBoSP) in NHS GG&C (April 2009 to April 2011) had previously been created. The screening algorithm and derivation of this cohort has been reported previously (Mansouri et al. 2013). All colonoscopists have to comply with strict quality control measures and require to have Joint Advisory Group (JAG) accreditation. In addition, all polyps identified are removed at colonoscopy.

A representative sample of 207 polyps from 134 individuals was chosen for inclusion in the study. All samples were processed in a single pathology department (Glasgow Royal Infirmary). All polyps were greater than 10mm in size at histological analysis. Details on site and macroscopic morphological appearance of the polyp were obtained from endoscopic reports. Details on post-fixation size, grade of dysplasia and microscopic appearance were obtained from pathology reports. Patient details included age, sex, and socioeconomic deprivation status. The Scottish Index of Multiple Deprivation (2009) was used as a measure of deprivation as has been described previously (SIMD 2009). Details on patient usage of Aspirin was obtained from pre-assessment documentation. Follow-up details on any further colonoscopies and details of recurrent or metachronous neoplasia (colorectal cancer or dysplastic polyp), were obtained from patient medical records on a case-by-case basis.

Ethical approval for use of this tissue was obtained from the West of Scotland Research Ethics Committee (12/WS/0152 – An investigation into tumour and host prognostic factors in early stage colorectal cancer and their correlation with clinical outcome. June 2012. Amendment approved December 2012).

### Immunohistochemistry

Assessment of inflammatory cell phenotype infiltrate was carried out by immunohistochemistry. Representative archival formalin fixed paraffin embedded tissue blocks were retrieved from archive and 2.5μm sections cut. Sections were then dewaxed rehydrated through graded alcohol. An autostainer (ThermoFisher, Autostainer 480s) was used to perform staining. Antigen retrieval was carried out in a PT module (ThermoFisher) using ThermoFisher dewax/retrieve solution pH9. Primary antibody was applied for 20 minutes at RT following antigen retrieval. Signal was amplified and visualised using the ThermoFisher Quanto kit and the diaminobenzidine (DAB) colour developer. Cell surface antigens were evaluated for T-lymphocytes (CD3+) dilution 1:300 (ThermoFisher), cytotoxic T-lymphocytes (CD8+) dilution 1:100 (ThermoFisher), helper T-lymphocytes (CD45+) dilution 1:500 (ThermoFisher) and macrophages (CD68+) dilution 1:5000 (ThermoFisher).

### Assessment of inflammatory infiltrate

All stained slides were converted to electronic format using a high resolution digital scanner (Hamamatsu NanoZoomer, Hamamatsu, Welwyn Garden City) and images viewed and assessed using Slidepath Digital Image Hub and Image Analysis module (Leica Microsystems, Wetzlar, Germany). The whole slide was then analysed in a semi-quantitative manner to assess intra-epithelial cell infiltrate at a resolution of 20x. Immune-cell infiltrate was graded on a four-point scale as absent, weak, moderate or strong (Supplementary Figures 1 to 4). Following initial scoring, this was further dichotomised into low (absent and weak) and high (moderate and strong) for the purpose of analysis. A total of 30 slides for each stain were scored independently by two observers to confirm consistency of scoring (DM and JHP). The remainder of the slides were then scored by a single observer (DM). The inter-observer intraclass coefficents for each subtype were: CD3+ = 0.66, CD8+ = 0.66, CD45+ =0.29(0.69 following retraining) and CD68+ = 0.79. A kappa value above 0.6 indicates good concordance.

A combined CD3+/CD8+ score was derived for each polyp to reflect the intra-epithelial densities of these two stains in manner similar to the recognised ‘Immunoscore’ for established tumours (Galon et al. 2014). This was defined as such: high combined CD3+/CD8+ score polyps were CD3+ high and CD8+ high, low combined CD3+/CD8+ score polyps were CD3+ low and CD8+ low, medium combined CD3+/CD8+ score were all other polyps.

Furthermore, the total inflammatory infiltrate was then derived for each polyp based on a combination of lymphocyte (CD3+) and macrophage (CD68+) scores. This was defined as such: high total inflammatory infiltrate polyps were CD3+ high and CD68+ high, low total inflammatory infiltrate polyps were CD3+ low and CD68+ low, medium total inflammatory infiltrate were all other polyps.

### Statistical Methods

Associations between categorical variables were examined using the χ2 test. For ordered variables with multiple categories the χ2 test for linear trend was used. Wilcoxon signed-rank test was used for analysis of paired variables. Binary logistic regression analysis was used to assess risk of neoplasia recurrence. A value of p<0.05 was considered statistically significant. Statistical analysis was performed using SPSS software (SPSS Inc., Chicago, IL, USA)

## Results

### Per polyp analysis

A total of 207 polyps from 134 patients were included. 107 were high grade (HGD) and 100 were low grade (LGD). The median age of patients was 65 years and 33 (25%) were female. The majority of polyps were left sided, pedunculated and were between 10mm and 20mm in size. Only 23 (11%) of patients reported aspirin use (Table 1). Comparing HGD and LGD polyps there were more older, female and less deprived patients in the HGD group (both p<0.05). HGD polyps were more likely to be larger and have a villous component (both p<0.05) (Table 1). Examining the inflammatory infiltrate in polyps, high levels of CD3+, CD8+, CD45+ and CD68+ were observed in 67%, 25%, 67% and 72% of cases respectively. CD3+ infiltrate was significantly higher in HGD polyps compared to LGD polyps (74% vs 69%, p<0.05). CD8+ infiltrate was significantly higher in HGD polyps compared to LGD polyps (36% vs 13%, p< 0.001) where as CD45+ infiltrate was not significantly different(69% vs 64%, p=0.401). The combined CD3+/CD8+ score was significantly higher in HGD polyps than LGD polyps (35% vs 11%, p<0.001). There was no significant difference in CD68+ infiltrate (74% vs 70%, p=0.540) (Table 2) or total inflammatory cell infiltrate (calculated from CD3+ and CD68+) (p=0.226).

**Table 1:** Baseline characteristics of adenomatous polyps by grade of dysplasia (per polyp analysis)

|  | All polyps<br>n(%) | Low-grade<br>polyps<br>n(%) | High-grade<br>polyps<br>n(%) | p-value |
| --- | --- | --- | --- | --- |
|  | 207 | 100 | 107 |  |
| **Age** |  |  |  |  |
| <65 | 111 (54) | 61 (61) | 50 (47) | 0.040 |
| ≥65 | 96 (46) | 39 (39) | 57 (53) |  |
| **Sex** |  |  |  |  |
| Male | 166 (80) | 88 (88) | 78 (73) | 0.006 |
| Female | 41 (20) | 12 (12) | 29 (27) |  |
| **Deprivation Quintile** |  |  |  |  |
| 1 (most deprived) | 81 (40) | 49 (51) | 32 (30) | 0.003 |
| 2 | 25 (12) | 8 (8) | 17 (16) |  |
| 3 | 36 (18) | 21 (21) | 15 (14) |  |
| 4 | 23 (11) | 6 (6) | 17 (16) |  |
| 5 (least deprived) | 39 (19) | 13 (13) | 26 (24) |  |
| **Aspirin exposure** |  |  |  |  |
| Yes | 23 (11) | 9 (9) | 14 (13) | 0.377 |
| No | 182 (89) | 89 (91) | 93 (87) |  |
| **Adenoma location** |  |  |  |  |
| Proximal to splenic flexure | 27 (13) | 16 (16) | 11 (10) | 0.223 |
| Distal to splenic flexure | 180 (87) | 84 (84) | 96 (90) |  |
| **Macroscopic appearance** |  |  |  |  |
| Sessile | 53 (26) | 28 (28) | 25 (23) | 0.445 |
| Pedunculated | 154 (74) | 72 (72) | 82 (77) |  |
| **Size (mm)** |  |  |  |  |
| 10-12 | 48 (23) | 32 (32) | 16 (15) | 0.002 |
| 13-15 | 54 (54) | 25 (25) | 29 (27) |  |
| 16-20 | 59 (59) | 28 (28) | 31 (29) |  |
| >20 | 46 (22) | 15 (15) | 31 (29) |  |
| **Microscopic appearance** |  |  |  |  |
| Tubular | 88 (43) | 50 (50) | 38 (36) | 0.036 |
| Tubulovillous/villous | 119 (57) | 50 (50) | 69 (64) |  |

**Table 2:** Comparison of inflammatory infiltrate by grade of adenomatous polyp (per polyp analysis)

|  | All polyps<br>n(%) | Low-grade<br>polyps<br>n(%) | High-grade<br>polyps<br>n(%) | p-value |
| --- | --- | --- | --- | --- |
|  | 207 | 100 | 107 |  |
| **CD3+** |  |  |  |  |
| High | 140 (67) | 61 (61) | 79 (74) | 0.049 |
| Low | 67 (32) | 39 (39) | 28 (26) |  |
| **CD8+** |  |  |  |  |
| High | 52 (25) | 13 (13) | 39 (36) | <0.001 |
| Low | 155 (75) | 87 (87) | 68 (64) |  |
| **CD45+** |  |  |  |  |
| High | 137 (67) | 63 (64) | 74 (69) | 0.401 |
| Low | 69 (33) | 36 (36) | 33 (31) |  |
| **CD68+** |  |  |  |  |
| High | 149 (72) | 70 (70) | 79 (74) | 0.540 |
| Low | 58 (28) | 30 (30) | 28 (26) |  |
| **Combined CD3+/CD8+ score** |  |  |  |  |
| Low | 64 (31) | 38 (38) | 26 (24) | <0.001 |
| Med | 94 (46) | 50 (51) | 44 (41) |  |
| High | 48 (23) | 11 (11) | 37 (35) |  |
| **Total inflammatory infiltrate** |  |  |  |  |
| Low | 30 (15) | 17 (17) | 13 (12) | 0.226 |
| Med | 65 (31) | 35 (35) | 30 (28) |  |
| High | 112 (54) | 48 (48) | 64 (60) |  |

Both patient and polyp related factors were then examined to identify features associated with altered inflammatory infiltrates. There was no difference in the degree of CD3+, CD8+, CD45+ or CD68+ inflammatory infiltrate with regards to patient factors such as age, sex or deprivation. Aspirin exposure was associated with a significantly higher level of CD45+ infiltrate (p=0.007). With regards to polyp factors, larger polyps were associated with a significantly higher level of CD8+ infiltrate (p=0.004) and polyps with a villous component had significantly higher levels of CD8+ infiltrate (p=0.021) Larger polyps (p=0.003) and polyps with a villous component (p=0.043) had higher combined CD3+/CD8+ scores. Total inflammatory infiltrate as calculated from CD3+ and CD68+ was not related to patient or polyp factors (Table 3).

**Table 3:**
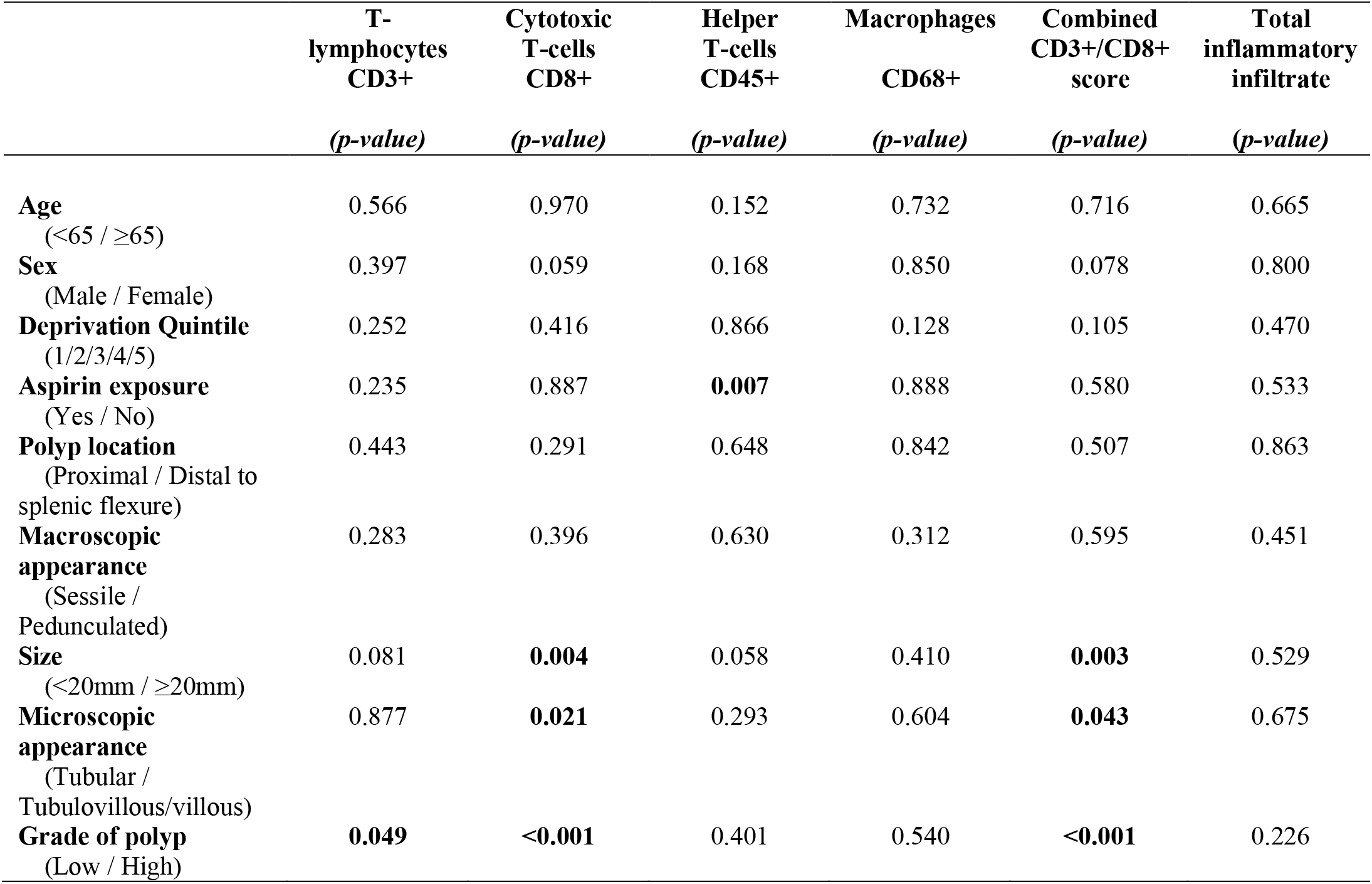
Associations between adenomatous polyp and host variables and degree of inflammatory infiltrate (per polyp analysis)

### Per patient analysis

In order to examine whether alterations in the microenvironment were related to altered host response, a per patient paired analysis was then carried out of those patients with multiple polyps (n=46 patients) (Figure 1). This included those with multiple low-grade dysplastic polyps (n=24 patients) and those with multiple mixed low and high-grade dysplastic polyps (n=20 patients). Due to low numbers, analysis of the 2 patients with multiple high-grade polyps was not carried out. On paired testing using Wilcoxon signed-rank analysis there was an increase in CD3+ (p=0.059), CD68+ (p=0.046) and total inflammatory infiltrate (p=0.021) in high-grade polyps of those who had both low and high-grade dysplasia. There was no change in CD8+ (p=0.705), CD45+ (p=0.605), or combined CD3+/CD8+ score (p=0.218). No significant changes in inflammatory infiltrate were seen between polyps in those patients with low grade dysplasia only (CD3+, p=0.317; CD8+, p=0.083; CD45+, p=0.206; CD68+, p=0.705; combined CD3+/CD8+ score, p=0.109; total inflammatory infiltrate, p=0.617) (Figures 2a & b).

**Figure 1:**
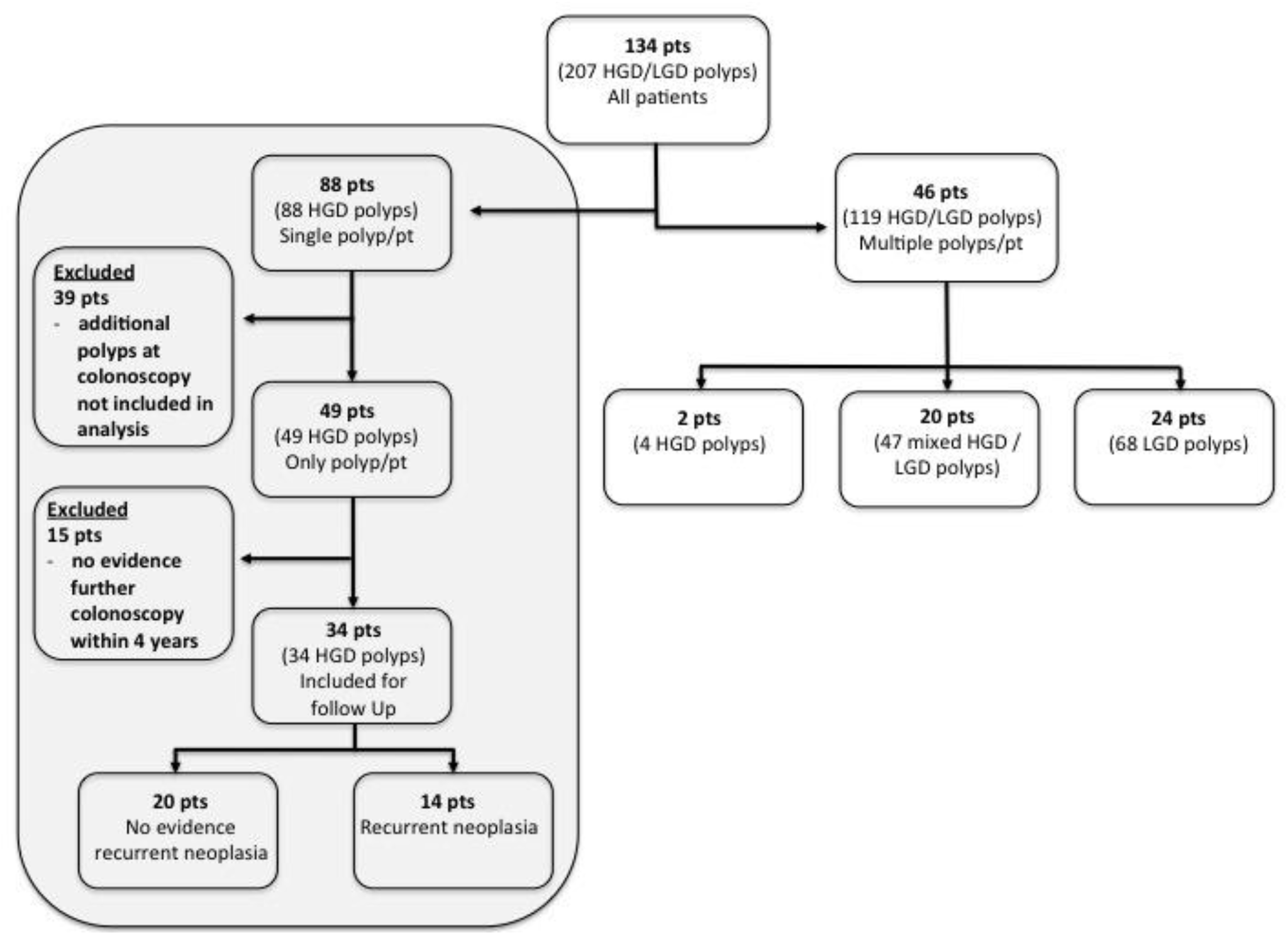
Outline of patient cohort and explanation of per patient analysis in those with multiple polyps, and follow up in those with solitary polyps (shaded box), (HGD = High-grade dysplasia, LGD = Low-grade dysplasia)

**Figure 2a:**
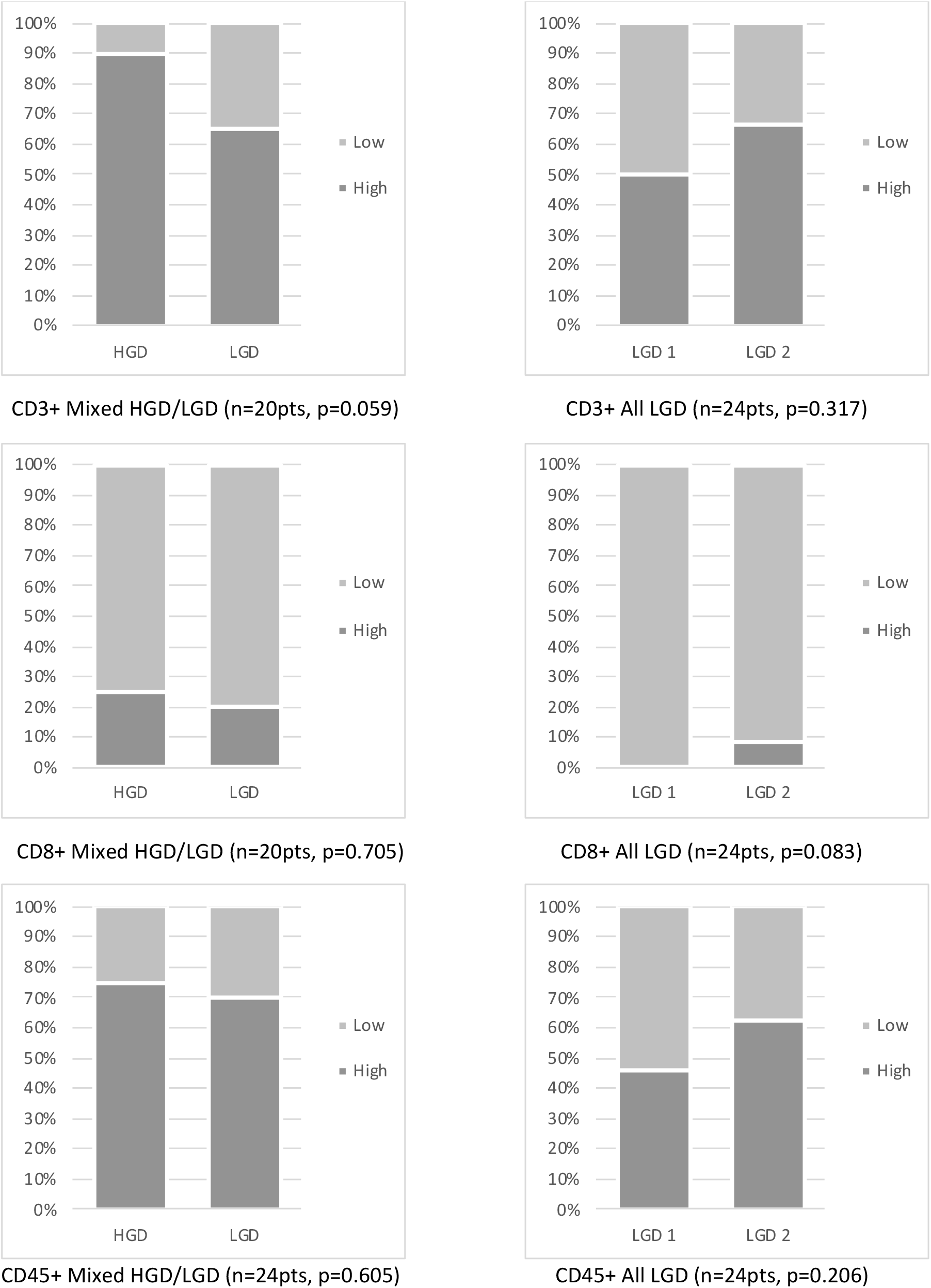
Per patient paired analysis of CD3+, CD8+, CD45+ T-lymphocyte cell infiltrate changes between polyps in patients with multiple polyps (mixed high/low grade dysplasia or low grade dysplasia only)

**Figure 2b:**
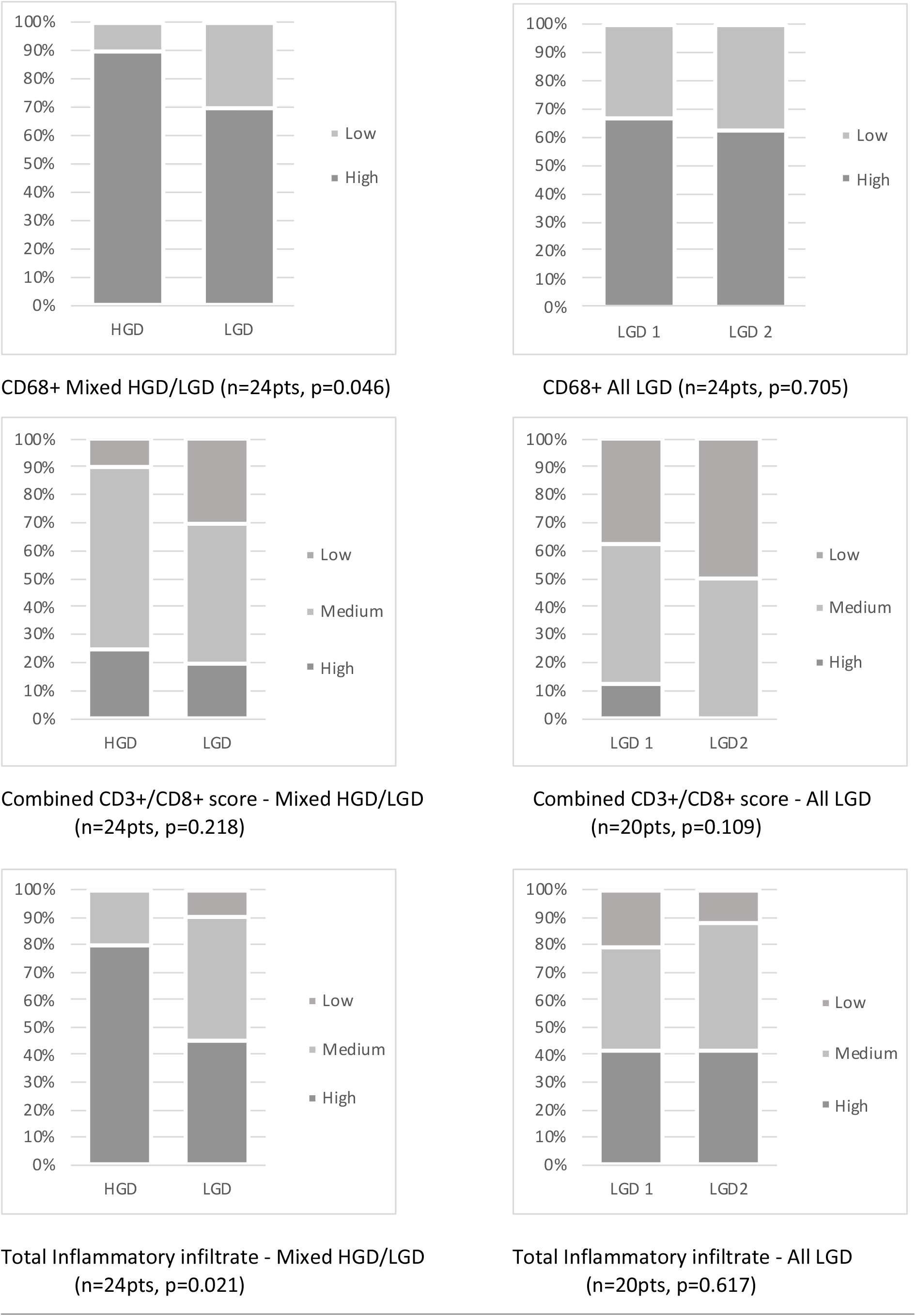
Per patient paired analysis of CD68+ macrophage, combined CD3+/CD8+ score and total inflammatory cell infiltrate changes between polyps in patients with multiple polyps (mixed high/low grade dysplasia or low grade dysplasia only)

### Risk of future / metachronous neoplasia

In those patients with a single polyp who had been included in the analysis (n=88 patients) outcome was assessed with regard to risk of metachronous neoplasia. Of the 88 patients, 39 (44%) patients were excluded as they had multiple polyps at index colonoscopy that had not been assessed in this analysis. On follow up, with a minimum of 4 years, 34 patients had undergone at least 1 colonoscopy whereby 14 had evidence of further neoplasia (Figure 1). There was no significant association between any of the measured local inflammatory infiltrate parameters and metachronous polyp (Table 4).

**Table 4:** Associations between inflammatory infiltrate and future / metachronous neoplasia in those with solitary polyps.

| Inflammatory parameter | Infiltrate | No future polyp<br>n(%) | Future polyp<br>n(%) | *p-value* |
| --- | --- | --- | --- | --- |
| CD3+ T lymphocytes | High | 15 (75) | 12 (86) | 0.672 |
|  | Low | 5 (25) | 2 (14) |  |
| CD8+ Cytotoxic T cells | High | 11 (58) | 7 (50) | 1.000 |
|  | Low | 9 (42) | 7 (50) |  |
| CD45+ Helper T cells | High | 13 (65) | 11 (78) | 0.467 |
|  | Low | 7 (35) | 3 (22) |  |
| CD68+ Macrophages | High | 18 (90) | 10 (70) | 0.202 |
|  | Low | 2 (10) | 4 (30) |  |
| Combined CD3+/CD8+ score | Low | 5 (25) | 2 (14) | 0.839 |
|  | Med | 4 (20) | 5 (36) |  |
|  | High | 11 (55) | 7 (50) |  |
| Total inflammatory infiltrate | Low | 2 (10) | 2 (15) | 0.746 |
|  | Med | 3 (15) | 2 (15) |  |
|  | High | 15 (75) | 10 (70) |  |

## Discussion

The present study shows that, within the context of a colorectal cancer screening programme, there was an increase in inflammatory cell infiltrate with progression from low-grade to high-grade dysplastic polyps. This was evident whether analysed within or between patients. Therefore, it appears that there is a specific host response to dysplastic changes in colorectal adenomas. Given the prognostic value of the tumour inflammatory cell infiltrate in established cancer (Roxburgh and McMillan 2012) it may be that the inflammatory cell infiltrate will inform the likely outcome in patients with dysplastic polyps.

The results of the present study are consistent with the observation that there is a specific interaction between adenomatous cells and the microenvironment (Cui, Yuan et al. 2009; McLean, Murray et al. 2011; Cui, Shi et al. 2012). A full understanding of the local inflammatory microenvironment of colorectal adenomas is essential if we are to learn about why some adenomas progress. The immunoediting hypothesis suggests that for neoplasia to develop there is an immune profile shift from immunosurveillance to immunosuppression (Koebel, Vermi et al. 2007). Based on this theory, adenomatous polyps represent a neoplastic lesion in the equilibrium phase and hence identifying which polyps are appear more likely to escape would be of considerable benefit (Dunn, Old et al. 2004). Indeed, the findings of the present study are in keeping with this model, and an increasing lymphocytic infiltrate as a marker of progression should be considered further. If such results were to be confirmed, then potential modulation of this local host inflammatory microenvironment to prevent escape could become of interest.

It is interesting to consider the finding of the present study with regards to patient outcome. Currently, follow-up of patients with adenomatous polyps is based on size, grade and number of polyps as previous studies have shown that patients with larger, high-grade and multiple polyps are at a higher risk of recurrence and of malignant transformation (Rutter et al. 2020). This surveillance paradigm has been in use in the UK for around two decades (Cairns et l. 2002, Cairns, Scholefield et al. 2010). However, this current risk stratification technique is far from ideal and a recent population based study has suggested that there is little benefit in terms of cancer incidence reduction in following these guidelines (Loberg, Kalager et al. 2014). With just under half of all those attending for a colonoscopy following a positive bowel screening test having adenomatous polyps detected, accurate prognostic stratification is vital if we are to avoid unnecessary follow-up colonoscopy in a large proportion of the population (Logan, Patnick et al. 2012). If such a link were to be proven, then it would represent a potential immunomodulatory target to help prevent the progression of adenomas in a pre-malignant phase.

The present study was predominantly cross-sectional in nature and lacked numbers to study outcome with sufficient power. However, a pilot group of 34 patients who had solitary polyps excised were followed up and a high rate of recurrent or metachronous polyps were noted (over 40% within 4 years). This should inform planning for future studies in larger numbers to explore this further and could also include those with multiple polyps. However, care should be taken with such analysis as it would be prone to potential confounding factors such as the heterogeneity of inflammatory infiltrate between polyps within the same person as has been demonstrated within the present study. Therefore, it may be useful to examine other polyp characteristics including gene expression and transcription patterns along with inflammatory infiltrate to identify additional factors associated with advanced adenomas and indeed metachronous polyp formation. Indeed, there is some existing evidence that such omics techniques can be applied to excised polyps in such a manner (Berger et al. 2018, Druliner et al. 2018, van Lanschot et al. 2019).

The strengths of the present study are that it is, to date, the largest study examining the local inflammatory response in colorectal adenomas. It has examined a variety of inflammatory cells using robust immunochemical techniques. In including a per patient analysis, changes in inflammatory infiltrate between different lesions within the same colon is achieved and adds to the robustness of the findings. In addition, it has included details on aspirin usage, a potential confounder that has been missing from previous studies (Cui, Yuan et al. 2009; McLean, Murray et al. 2011). A potential additional weakness is the use of CD68+ as a marker for macrophage infiltration. When considering the immune microenvironment, macrophage subtypes associated with either an increase in the adaptive or the innate response can be of interest and CD68+ staining does not account for that. Further studies examining macrophage subtypes to explore this concept are planned in this cohort. One further potential weakness may be our use of a derived total inflammatory infiltrate rather than a true observed one. For example, in colorectal cancer the Klintrup-Makinen score, assessing inflammatory cell infiltrate at the invasive margin on routinely stained H&E slides, has been widely validated as means of observing a total inflammatory cell infiltrate and has been correlated with patient outcome (Klintrup, Makinen et al. 2005). However, given the nature of endoscopic polyp resections, where there is no clear margin in a large proportion of cases, means that inflammatory cell infiltrate can only be examined within the adenoma itself and not at the margin. Therefore, the Klintrup-Makinen score cannot accurately be assessed, however recent findings in colorectal cancer suggest that the lymphocyte subtype and location of the infiltrate is less prognostically important than the density (Alexander et al. 2020, Haruki et al. 2020). The lack of margin assessment also the limited the use of the well validated Immunoscore as described by Galon et al. (Galon et al. 2014). However, the use of a combined intra-epithelial CD3+/CD8+ score has been proposed here as a method of compensating for this and further work in larger numbers is planned utilising this adaptation. Moreover, studies have shown that it is in fact density, rather than location that is the most important aspect of inflammatory infiltrate related to outcome when examined in established disease (Alexander et al. 2020).

In conclusion, the present study has shown an increase in inflammatory infiltrate with progression from low-grade to high-grade dysplasia. This would suggest a specific response to early disease progression confirming increased host immunosurveillance. Such a finding may have a use in the prognostic stratification and treatment of dysplastic polyps, and as a mechanism of predicting the likelihood of future polyps given the known associations between advanced features and metachronous adenomas.

## Data Availability

Access to study data available on request.

## Acknowledgements

This work was funded by The Cunningham Trust, St Andrews, Scotland (Scottish Charity number SCO13499, Ref ACC/KWF/CT12/21, awarded 23/11/2012.

## Disclosures

None

## Funding

The Cunningham Trust, SCO13499, Ref ACC/KWF/CT12/21

## Notes

### Competing Interest Statement

The authors have declared no competing interest.

### Funding Statement

Study funded by a research grant from charitable organisation The Cunningham Trust, SCO13499, Ref ACC/KWF/CT12/21.

### Author Declarations

Ethical approval for use of this tissue was obtained from the West of Scotland Research Ethics Committee (12/WS/0152 An investigation into tumour and host prognostic factors in early stage colorectal cancer and their correlation with clinical outcome. June 2012. Amendment approved December 2012).

